# Evolution of COVID-19 mortality over time: results from the Swiss hospital surveillance system (CH-SUR)

**DOI:** 10.1101/2021.09.14.21263153

**Authors:** Maroussia Roelens, Alexis Martin, Brian Friker, Filipe Maximiano Sousa, Amaury Thiabaud, Beatriz Vidondo, Valentin Buchter, Céline Gardiol, Jasmin Vonlanthen, Carlo Balmelli, Manuel Battegay, Christoph Berger, Michael Buettcher, Alexia Cusini, Domenica Flury, Ulrich Heininger, Anita Niederer-Loher, Thomas Riedel, Peter W. Schreiber, Rami Sommerstein, Nicolas Troillet, Sarah Tschudin-Sutter, Pauline Vetter, Sara Bernhard-Stirnemann, Natascia Corti, Roman Gaudenz, Jonas Marschall, Yvonne Nussbaumer-Ochsner, Laurence Senn, Danielle Vuichard-Gysin, Petra Zimmermann, Franziska Zucol, Anne Iten, Olivia Keiser, CH-SUR study group

## Abstract

**Background:** When comparing the periods of time during and after the first wave of the ongoing SARS-CoV-2/COVID-19 pandemic in Europe, the associated COVID-19 mortality seems to have decreased substantially. Various factors could explain this trend, including changes in demographic characteristics of infected persons, and the improvement of case management. To date, no study has been performed to investigate the evolution of COVID-19 in-hospital mortality in Switzerland, while also accounting for risk factors.

**Methods:** We investigated the trends in COVID-19 related mortality (in-hospital and in-intermediate/intensive-care) over time in Switzerland, from February 2020 to May 2021, comparing in particular the first and the second wave. We used data from the COVID-19 Hospital-based Surveillance (CH-SUR) database. We performed survival analyses adjusting for well-known risk factors of COVID-19 mortality (age, sex and comorbidities) and accounting for competing risk.

**Results:** Our analysis included 16,030 episodes recorded in CH-SUR, with 2,320 reported deaths due to COVID-19 (13.0% of included episodes). We found that overall in-hospital mortality was lower during the second wave of COVID-19 compared to the first wave (HR 0.71, 95% CI 0.69 – 0.72, p-value < 0.001), a decrease apparently not explained by changes in demographic characteristics of patients. In contrast, mortality in intermediate and intensive care significantly increased in the second wave compared to the first wave (HR 1.48, 95% CI 1.42 – 1.55, p-value < 0.001), with significant changes in the course of hospitalisation between the first and the second wave.

**Conclusion:** We found that, in Switzerland, COVID-19 mortality decreased among hospitalised persons, whereas it increased among patients admitted to intermediate or intensive care, when comparing the second wave to the first wave. We put our findings in perspective with changes over time in case management, treatment strategy, hospital burden and non-pharmaceutical interventions. Further analyses of the potential effect of virus variants and of vaccination on mortality would be crucial to have a complete overview of COVID-19 mortality trends throughout the different phases of the pandemic.

## Introduction

Many countries around the world, and in particular in Europe, experienced an evolution of the COVID-19 pandemic over time with a first wave occurring in Spring 2020, followed by a substantial decrease in the number of COVID-19 cases during the intermediate period, and then by the emergence of a second wave in Fall 2020. The chronology of the different COVID-19 waves is tightly related to governments’ responses to the epidemic, which varied highly over time and country, often tightening and relaxing non-pharmaceutical interventions (hereafter NPI, i.e. infection control measures including lockdown, travel banning and the setup of testing and tracing programs) depending on the number of diagnosed COVID-19 cases.

The assessment of COVID-19 associated mortality is crucial not only to evaluate the impact of the pandemic, but also to evaluate the improvement of knowledge on the disease and of patient care, as well as the effectiveness of measures adopted to control the outbreak. It has been hypothesized that COVID-19 mortality might have been lower after the first wave in Europe [1–3].

Crude calculations from number of cases and deaths reported by the Federal Office of Public Health (FOPH) [4, 5] yield case fatality ratios (CFR, i.e. proportion of deaths among all infected persons identified) for COVID-19 in Switzerland and Liechtenstein of approximately 5.4% between February and end of April 2020 ; 0.89% between May and September 2020 and 1.6% from October 2020 to mid-February 2021. This change in mortality could potentially be explained by several factors. First, the restrictions in COVID-19 testing during the first wave likely resulted in a high CFR, whereas the increase in testing capacity over time and especially since November 2020 could decrease the CFR estimate over time. However, since testing restrictions were not applied to hospitalised persons, it should not affect the in-hospital mortality rate (i.e. proportion of deaths among all persons hospitalised with COVID-19 diagnosis). Then, it could relate to changes in demographic characteristics of the population. In particular, older age and comorbidities are very well-known risk factors for mortality of SARS-CoV-2 infected persons [6–9]. Moreover, earlier access to medical care and improved care in hospitals could also contribute to a lower morbidity and mortality over time. Some studies showed an improvement of survival among critical care patients with COVID-19 [10], likely due to improvements in treatment strategies and management of severe cases.

To our knowledge, no study has been performed to investigate the evolution of COVID-19 related morbidity and mortality among hospitalized patients at a national scale in Switzerland, while also accounting for risk factors. Therefore, the aim of the present study is to investigate the trends in COVID-19 related mortality over time in Switzerland, comparing in particular the first and the second wave, using data from the COVID-19 Hospital-based Surveillance (CH-SUR) database [11].

## Materials and Methods

### Data source

CH-SUR (COVID-19 Hospital Based Surveillance) is a prospective hospital surveillance system for COVID-19 coordinated by the Swiss Federal Office of Public Health (FOPH) and the Institute of Global Health (ISG) of the University of Geneva [11, 12], designed to capture detailed information on COVID-19 hospitalised patients in Switzerland. It includes patients hospitalised for more than 24h and diagnosed with COVID-19 confirmed by a laboratory test (e.g. polymerase chain-reaction test, PCR), plus those confirmed by a serology or with a CT-scan and thorax radiography compatible with COVID-19 diagnosis since November 14^th^ 2020. To date, 21 Swiss hospitals are participating in that surveillance (including university hospitals and cantonal hospitals), and are representative of the overall COVID-19 situation in Switzerland [9].

For each COVID-19 episode recorded in CH-SUR, various information on demographics, admission, clinical information (including comorbidities, complications, admission to intermediate care unit and/or intensive care unit -hereafter IMCU/ICU- and treatments) and follow-up (death, discharge or transfer) are collected. Further information on CH-SUR can be found elsewhere [11]. Of note, since CH-SUR was setup first for surveillance purposes, in a period of high burden for participating hospitals, priority was given to collection of data on admission and follow-up, to enable timely estimates of numbers regarding new cases and outcomes. Hence, the documentation of additional complementary information (which is relatively time consuming) is optional, and is often provided with some delay compared to the mandatory information on admission and follow-up. At the time of data export, the clinical complementary information was complete for about 86% of recorded episodes.

For that analysis, we included all adult (age ≥ 18 years) patients in CH-SUR diagnosed between February 26st 2020 and May 15th 2021, as recorded by June 21st 2021. Patients diagnosed after May 15^th^ were excluded to reduce the effect of incomplete cases with yet unknown outcome. Out of the 21 hospitals participating to CH-SUR, one hospital had too incomplete data to be included in our analysis (clinical complementary information provided for less than 50% of episodes), and 6 were paediatric hospitals (hence reporting no adult case), consequently the data we used was collected from only 14 hospitals.

### Statistical analysis

We investigated four different time periods in our analyses, corresponding to the different waves of COVID-19 observed in Switzerland, which are defined in Table 1. However, due to the limited time span of the last period and the partial incompleteness of these data, the interpretation of results focus on the comparison between the first and the second wave.The outcome of a COVID-19 episode was either death due to COVID-19, death due to other cause than COVID-19, discharge (including both recovered patients who go home and those discharged -presumably not infectious anymore- to a nursing home) or transfer to a hospital outside the surveillance network. Patients with no outcome documented were assumed to be still in care.

**Table 1:** Definition of time periods for COVID-19 waves in Switzerland

| Time period | Start date | End date |
| --- | --- | --- |
| First wave | 2020-02-01 | 2020-04-30 |
| Intermediate phase | 2020-05-01 | 2020-09-30 |
| Second wave | 2020-10-01 | 2021-02-14 |
| Last period | 2021-02-15 | 2021-06-21<br>(date of data extraction) |

We conducted a survival analysis, for which time-to-event was calculated from the date of the first COVID-19 diagnosis (i.e. date of first sample positive to SARS-CoV-2 infection or date of first clinical diagnosis with imaging) to either the date of outcome or the censoring date. We accounted for competing risks, with death due to COVID-19 being the outcome of interest, and death due to other causes, discharge and transfer being considered as competing risks. Patients still in care were censored at the date of the data export (June 21st 2021) [13].

We calculated crude cumulative incidence functions (CIF) for death, overall and per time period, using the R package mstate [13, 14]. We used univariable and multivariable Fine and Gray models [15] to determine risk factors of mortality, adjusting for sex (male, female), age (as a continuous variable with restricted cubic splines [16]), time period of COVID-19 diagnosis (first wave, intermediate phase, second wave, last period), obesity (no, yes), smoking (no, yes), chronic respiratory disease (no, yes), cardiovascular disease (no, yes), renal disease (no, yes), oncological pathologies (no, yes), dementia (no, yes), immunosuppression (no, yes). We excluded patients with missing age, sex, or outcome date when the outcome was documented. We imputed missing values of all other covariables using multiple imputation by chained equation (R package mice [17]), adding the outcome and time to event to improve the results of the imputation. We ran the model on 20 imputed datasets for each analysis, and combined the estimates with Rubin’s rule [17]. For patients with multiple hospitalisations recorded in CH-SUR, we used the information on comorbidities provided at the first admission. We considered only the outcome of the last hospitalisation, and time-to-event was defined as the time from the date of first COVID-19 diagnosis to the date of the last outcome (or the time of censoring if considered still in care).

In a sensitivity analysis, we recalculated univariable and multivariable Fine and Gray models without multiple imputation, excluding all episodes with incomplete information on any of the adjusted covariables.

### Patients on intensive and intermediate care

Similar analyses were conducted for the subgroup of patients who have ever been admitted to intermediate care or ICU (hereafter IMCU/ICU) during their COVID-19 episode(s), with time-to-event being defined as the time from first admission in IMCU/ICU to either the date of outcome or the censoring date. Risk factors of mortality among that IMCU/ICU subgroup were explored using Fine and Gray models as detailed above, adding to the adjusted covariables: use of invasive ventilation during any of IMCU/ICU stays (no, yes), use of non-invasive ventilation during any of the IMCU/ICU stays (no, yes). For sensitivity purposes, we repeated this analysis but included only patients ever admitted to ICU.

## Results

### Patients and COVID-19 episodes characteristics

As of June 21st 2021, 16,513 COVID-19 episodes were recorded in CH-SUR, 16,030 of which were included in our analysis with 2,320 reported deaths (14.5% of included episodes), 2,084 of which being reported to be due to COVID-19 (13.0% of included episodes).

Characteristics of included episodes, overall and for the first and second waves of COVID-19, are shown in Table 2 and Table 3. The complete table of characteristics of all the included episodes, overall and stratified by time period, can be found in Appendix 1.

**Table 2:** Distribution of outcomes for the included COVID-19 episodes. P-value corresponds to comparison between the first wave and the second wave, from Chi-Square test.

|  | All<br>N=16030 | First wave<br>N=2781 | Second wave<br>N=10160 | P-value<br>1st wave/2nd wave |
| --- | --- | --- | --- | --- |
| Censored | 655 (4.1%) | 6 (0.2%) | 262 (2.6%) | P < 0.001 |
| Deceased – COVID-19 | 2084 (13%) | 434 (15.6%) | 1422 (14%) |  |
| Deceased - other | 236 (1.5%) | 10 (0.4%) | 198 (1.9%) |  |
| Discharged | 12569 (78.4%) | 2216 (79.7%) | 7986 (78.6%) |  |
| Transferred | 486 (3%) | 115 (4.1%) | 292 (2.9%) |  |

**Table 3:** Characteristics of COVID-19 episodes. For continuous variables, the values given are median (IQR). For categorical variables, percentages provided are calculated with respect to total number of episodes. P-values correspond to comparison between the first wave and the second wave, from Kruskal-Wallis Rank-Sum test for continuous variables and Chi-Square test for categorical variables.

|  |  | All<br>N=16030 | First wave<br>N=2781 | Second wave<br>N=10160 | P-value<br>1st wave/2nd wave |
| --- | --- | --- | --- | --- | --- |
| Time from diagnosis to outcome or censoring (d) |  | 13 (7 - 21) | 11 (6 - 21) | 13 (7 - 21) | P < 0.001 |
| Time from symptoms onset to diagnosis (d) |  | 2 (0 - 5) | 4 (1 - 7) | 2 (0 - 4) | P < 0.001 |
| Time from hospital admission to outcome (d) |  | 9 (5 - 19) | 10 (5 - 22) | 10 (5 - 19) | P = 0.29 |
| Time from diagnosis to hospital admission (d) |  | 0 (0 - 4) | 0 (0 - 2) | 0 (0 - 4) | P < 0.001 |
| Age (years) |  | 71 (58 - 81) | 69 (55 - 80) | 73 (61 - 82) | P < 0.001 |
| Sex | female | 6768 (42.2%) | 1103 (39.7%) | 4314 (42.5%) | P = 0.0086 |
|  | male | 9262 (57.8%) | 1678 (60.3%) | 5846 (57.5%) |  |
| Obesity | no | 9372 (58.5%) | 1496 (53.8%) | 6210 (61.1%) | P < 0.001 |
|  | yes | 4139 (25.8%) | 612 (22%) | 2626 (25.8%) |  |
|  | unknown | 2519 (15.7%) | 673 (24.2%) | 1324 (13%) |  |
| Smoking | no | 8573 (53.5%) | 1649 (59.3%) | 5310 (52.3%) | P < 0.001 |
|  | yes | 1578 (9.8%) | 230 (8.3%) | 1037 (10.2%) |  |
|  | unknown | 5879 (36.7%) | 902 (32.4%) | 3813 (37.5%) |  |
| Renal disease | no | 12317 (76.8%) | 2308 (83%) | 7607 (74.9%) | P < 0.001 |
|  | yes | 3022 (18.9%) | 462 (16.6%) | 2203 (21.7%) |  |
|  | unknown | 691 (4.3%) | 11 (0.4%) | 350 (3.4%) |  |
| Oncological pathology | no | 13419 (83.7%) | 2485 (89.4%) | 8449 (83.2%) | P < 0.001 |
|  | yes | 1909 (11.9%) | 285 (10.2%) | 1350 (13.3%) |  |
|  | unknown | 702 (4.4%) | 11 (0.4%) | 361 (3.6%) |  |
| Chronic respiratory disease | no | 12239 (76.4%) | 2159 (77.6%) | 7785 (76.6%) | P < 0.001 |
|  | yes | 3102 (19.4%) | 609 (21.9%) | 2026 (19.9%) |  |
|  | unknown | 689 (4.3%) | 13 (0.5%) | 349 (3.4%) |  |
| Cardiovascular disease | no | 9980 (62.3%) | 1902 (68.4%) | 5995 (59%) | P < 0.001 |
|  | yes | 5369 (33.5%) | 872 (31.4%) | 3824 (37.6%) |  |
|  | unknown | 681 (4.2%) | 7 (0.3%) | 341 (3.4%) |  |
| Dementia | no | 13939 (87%) | 2579 (92.7%) | 8721 (85.8%) | P < 0.001 |
|  | yes | 1372 (8.6%) | 186 (6.7%) | 1071 (10.5%) |  |
|  | unknown | 719 (4.5%) | 16 (0.6%) | 368 (3.6%) |  |
| Immunosuppression | no | 14020 (87.5%) | 2525 (90.8%) | 8935 (87.9%) | P < 0.001 |
|  | yes | 1272 (7.9%) | 214 (7.7%) | 863 (8.5%) |  |
|  | unknown | 738 (4.6%) | 42 (1.5%) | 362 (3.6%) |  |
| Any comorbidity | no | 2531 (15.8%) | 565 (20.3%) | 1294 (12.7%) | P < 0.001 |
|  | yes | 12853 (80.2%) | 2209 (79.4%) | 8543 (84.1%) |  |
|  | unknown | 646 (4%) | 7 (0.3%) | 323 (3.2%) |  |
| CURB-65 score | 0 | 4144 (25.9%) | 854 (30.7%) | 2130 (21%) | P < 0.001 |
|  | 1 | 5880 (36.7%) | 938 (33.7%) | 3841 (37.8%) |  |
|  | 2 | 4155 (25.9%) | 665 (23.9%) | 2921 (28.8%) |  |
|  | 3 | 1484 (9.3%) | 266 (9.6%) | 1019 (10%) |  |
|  | 4 | 306 (1.9%) | 53 (1.9%) | 219 (2.2%) |  |
|  | 5 | 31 (0.2%) | 5 (0.2%) | 20 (0.2%) |  |
|  | unknown | 30 (0.2%) | 0 (0%) | 10 (0.1%) |  |
| Ever admitted to ICU | no | 12720 (79.4%) | 2246 (80.8%) | 8355 (82.2%) | P < 0.001 |
|  | yes | 2483 (15.5%) | 527 (19%) | 1469 (14.5%) |  |
|  | unknown | 827 (5.2%) | 8 (0.3%) | 336 (3.3%) |  |
| Ever admitted to IMCU/ICU | no | 11775 (73.5%) | 1961 (70.5%) | 7832 (77.1%) | P < 0.001 |
|  | yes | 3455 (21.6%) | 812 (29.2%) | 1993 (19.6%) |  |
|  | unknown | 800 (5%) | 8 (0.3%) | 335 (3.3%) |  |

Of 2,084 deaths due to COVID-19 recorded in CH-SUR by the date of data export, 434 patients were diagnosed during the first wave, and 1,422 during the second wave. Crude in-hospital COVID-19 mortality ratios were 15.6% for patients diagnosed during the first wave, and 14.0% for those diagnosed during the second wave. Hospitalised COVID-19 patients were slightly older in the second wave (median 73 years, IQR 61 – 82 years) than in the first wave (median 69 years, IQR 55 – 80 years), and the proportion of male patients was slightly higher in the first wave (60.3%) than in the second wave (57.5%). Mild severity score at admission (CURB-65 score) tended to be more frequent in the second wave (21.0% with score 0, 37.8% with score 1, 28.8% with score 2) than in the first wave (30.7% with score 0, 33.7% with score 1, 23.9% with score 2).

Because the clinical complementary information is often entered in the database with some delay, data on comorbidities was slightly less complete for the second compared to the first wave (respectively 3.2% and 0.3% of episodes with missing information on presence of any comorbidity). However, hospitalised patients diagnosed during the second wave were more comorbid than those diagnosed during the first wave (resp. 86.8% and 79.6% of patients who had information on presence or absence of comorbidities available).

The median time from first COVID-19 diagnosis to outcome (or censoring) was 11 days (6 – 21 days) for the first wave, and 13 days (IQR 7 – 21 days) for the second wave. The duration of hospital stay was similar between the first (median of 10 days, IQR 5 – 22 days) and the second waves (median 10 days, IQR 5 - 19 days). Time from diagnosis to first hospital admission was often equal to zero in both waves, meaning that most patients are tested at hospital entry, however it tends to be shorter in the first wave (3^rd^ quartile 2 days) than in the second wave (3^rd^ quartile 4 days).

### Differences in overall in-hospital COVID-19 mortality by time period

Figure 1 shows cumulative incidence functions of death due to COVID-19 for each time period. Similar figures for the other outcomes (discharge, transfer, death due to other causes) can be found in Appendix 2. For the first wave, about 75% of deaths occurred within 15 days after diagnosis, and the probability of death increased from 5.1% after one week of diagnosis to 10.7% after 2 weeks, 14.1% after 4 weeks and 15.1% after 6 weeks. In contrast, for the second wave, about 67% of deaths occurred within 15 days after diagnosis, and the probability of death increased from 3.6% after one week to 8.4% after 2 weeks, 12.4% after 4 weeks and 13.5% after 6 weeks. This indicates that the probability of death was lower in the second wave than in the first wave, and that death tended to occur later after diagnosis in the second wave compared to the first wave.

**Figure 1:**
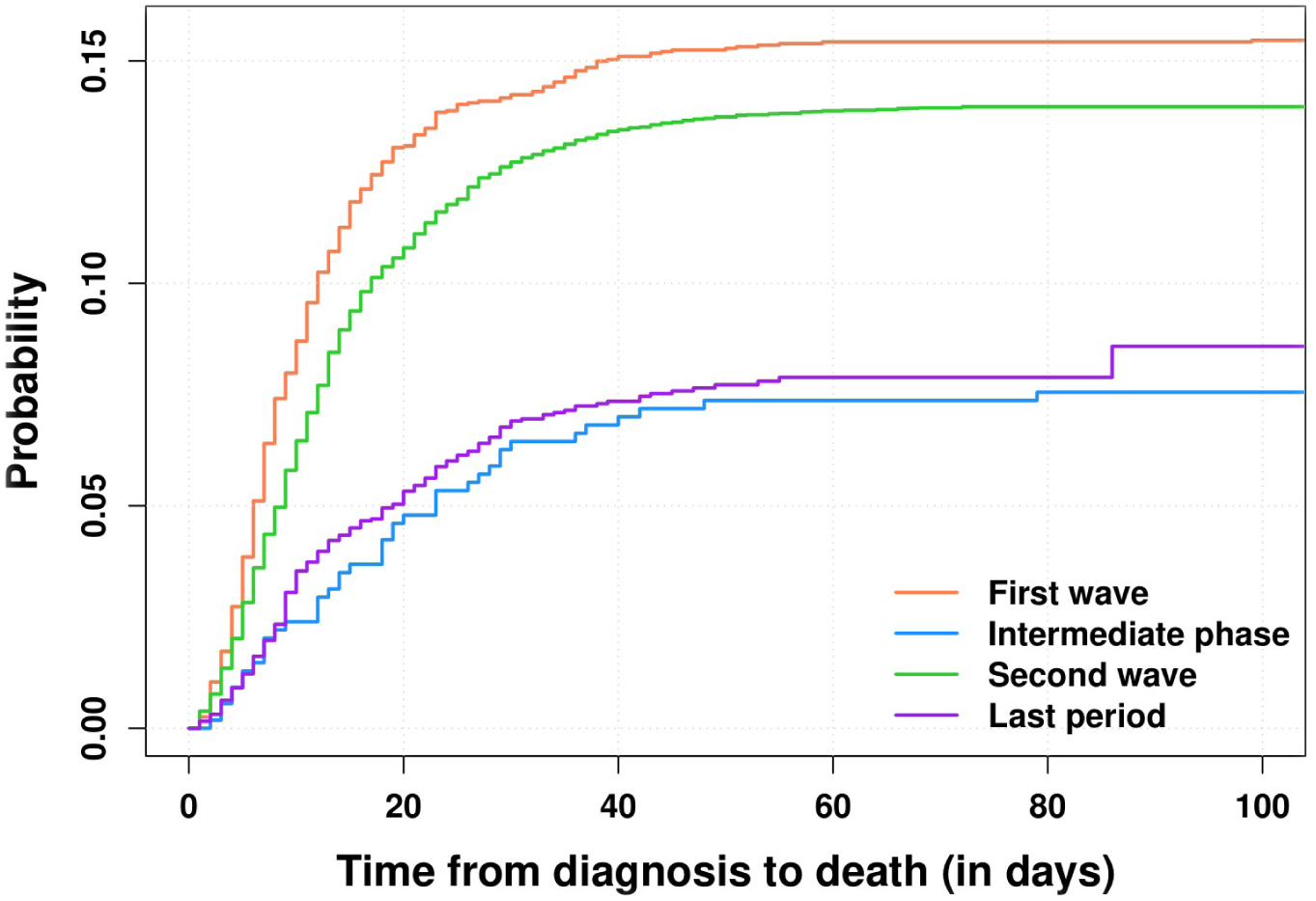
Cumulative incidences of death due to COVID-19 in CH-SUR, for the different waves of COVID-19.

Table 4 shows the hazard ratios (HRs) for in-hospital mortality for the different time periods, with 95% confidence intervals (95% CI), from the Fine and Gray models. HRs and 95% CI for all the covariables included in the models can be found in Appendix 3. The overall risk of death was lower for hospitalised patients diagnosed during the second wave compared to those diagnosed during the first wave (HR 0.71, 95% CI 0.69 – 0.73).

**Table 4:** Estimates of the effect of time period of diagnosis on mortality, from Fine and Gray models

|  | Multivariable, imputed | Multivariable, complete case | Univariable, complete case |
| --- | --- | --- | --- |
| Time period | P < 0.001 | P < 0.001 | P < 0.001 |
| First wave | 1.0 (ref) | 1.0 (ref) | 1.0 (ref) |
| Intermediate | 0.522 (0.486 – 0.561) | 0.652 (0.439 – 0.966) | 0.458 (0.333 – 0.631) |
| Second wave | 0.707 (0.69 – 0.725) | 0.72 (0.619 – 0.837) | 0.88 (0.79 – 0.98) |
| Last period | 0.595 (0.573 – 0.618) | 0.659 (0.52 – 0.835) | 0.459 (0.387 – 0.545) |

### Analysis of patients admitted to IMCU/ICU

As of June 21st 2021, 3,437 patients were admitted at least once to IMCU/ICU. For the 3,437 patients, 3,455 episodes were recorded (out of 16,030 included in the main analysis, i.e. 25.1%), 812 in the first wave and 1,993 in the second wave. Overall, for 3,429 of them the time to event variable was available (811 for the first wave, 1,980 for the second wave), with time to event being defined as time from first IMCU/ICU admission to outcome (or censoring). Characteristics of all those episodes, overall and stratified by time period of COVID-19 diagnosis can be found in Appendix 4. The proportion of patients ever admitted to IMCU/ICU was lower in the second wave than in the first wave (Table 3). The time elapsed from COVID-19 diagnosis to first admission into IMCU/ICU was longer in the second wave (median 4 days, IQR 0 – 8 days) than in the first wave (median 2 days, IQR 0 – 5 days). The time from first IMCU/ICU admission to outcome (or census) was similar in the second wave (median 16 days, IQR 9 – 30 days) and in the first wave (median 16 days, IQR 8 – 34 days). Moreover, the proportion of patients who ever went to IMCU/ICU and experienced invasive ventilation was higher in the first wave (53.6%) compared to the second wave (44.7%).

Similarly to all hospitalized patients, IMCU/ICU patients of the second wave were more comorbid than those of the first wave (88.4% versus 82.2%). Crude in-hospital COVID-19 mortality rates among patients who were admitted to IMCU/ICU are 19.0% and 27.1% for patients hospitalised during the first wave, and second wave, respectively.

Cumulative incidence functions for death due to COVID-19 during the different waves, limited to patients ever admitted to IMCU/ICU, are shown in Figure 2. Similar CIFs for the other outcomes (death due to other causes, discharge and transfer) can be found in Appendix 5. During the first wave, about 82% of deaths occurred within 4 weeks after first admission to IMCU/ICU; the probability of death among IMCU/ICU patients increased from 5.8% after one week to 11.2% after 2 weeks, 15.5% after 4 weeks and 17.9% after 6 weeks. During the second wave, about 88% of deaths occurred within 4 weeks after first admission to IMCU/ICU; the probability of death increased from 8.2% after one week to 14.6% after 2 weeks, 23.5% after 4 weeks and 25.9% after 6 weeks. This indicates that the probability of death among IMCU/ICU patients was higher in the second wave than in the first wave, and that death tended to occur earlier after IMCU/ICU admission in the second wave compared to the first wave.

**Figure 2:**
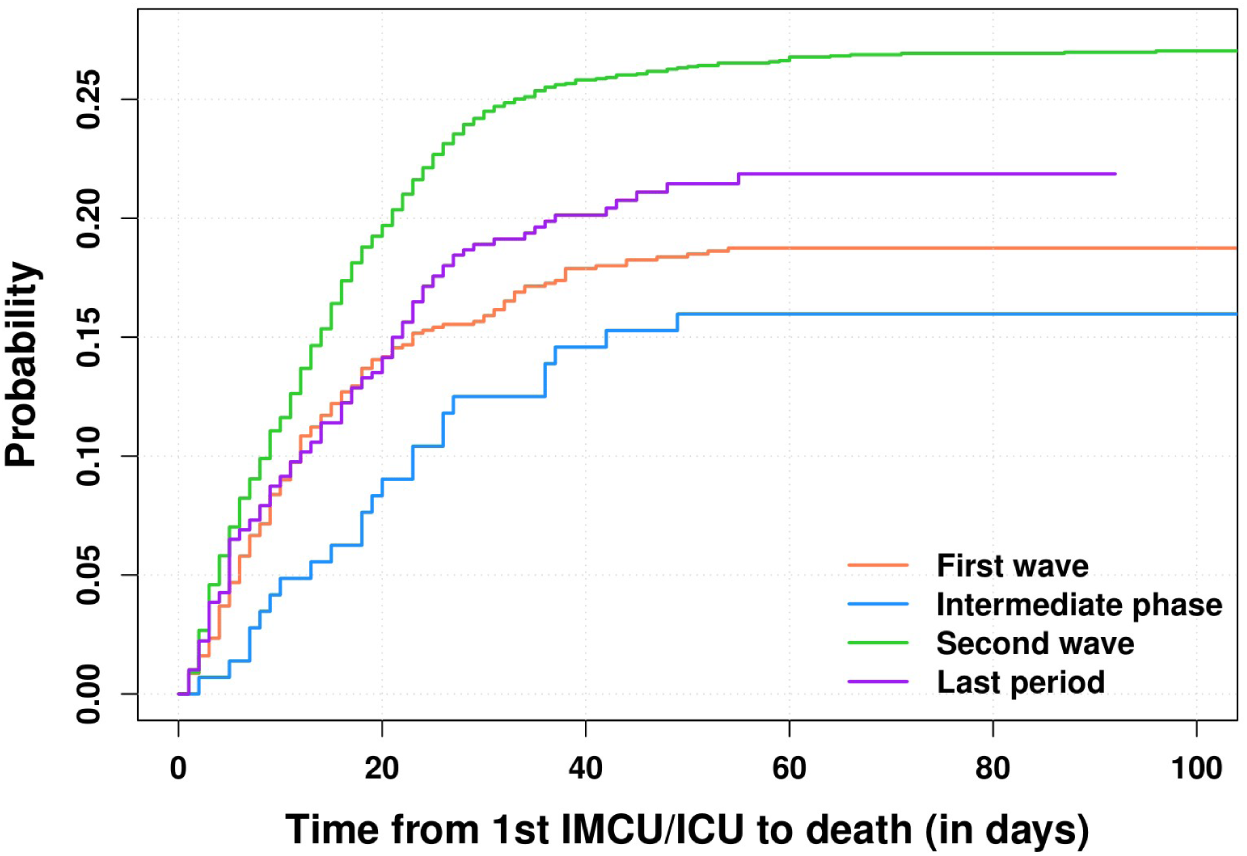
Cumulative incidence functions of death due to COVID-19 for patients admitted to IMCU/ICU, for the different time periods.

HRs associated with time period and use of invasive ventilation from Fine and Gray models are shown in Table 5. HRs and 95% CI for all the covariables included in the models can be found in Appendix 6. The risk of death was higher for patients who ever were admitted to IMCU/ICU during the second wave compared to the first wave (HR 1.49, 95% CI 1.43 – 1.55), and overall patients who underwent invasive ventilation also presented a higher mortality (HR 2.66, 95% CI 2.57 – 2.77), as well as patients who ever underwent non-invasive ventilation (HR 2.62, 95% CI 2.34 – 2.93).

**Table 5:** Estimates of the effect of time period and invasive ventilation on mortality among patients who ever went to IMCU/ICU, from Fine and Gray models.

|  |  | Multivariable, imputed | Multivariable, complete case | Univariable, complete case |
| --- | --- | --- | --- | --- |
| Time period |  | P < 0.001 | P = 0.039 | P < 0.001 |
|  | First wave | 1.0 (ref) | 1.0 (ref) | 1.0 (ref) |
|  | Intermediate | 0.946 (0.857 – 1.04) | 1 (0.414 – 2.44) | 0.808 (0.521 – 1.25) |
|  | Second wave | 1.49 (1.43 – 1.55) | 1.68 (1.12 – 2.52) | 1.5 (1.25 – 1.79) |
|  | Last period | 1.49 (1.41 – 1.58) | 1.85 (1.1 – 3.1) | 1.11 (0.867 – 1.43) |
| Invasive ventilation use |  | P < 0.001 | P = 0.0042 | P < 0.001 |
|  | No | 1.0 (ref) | 1.0 (ref) | 1.0 (ref) |
|  | Yes | 2.66 (2.57 – 2.77) | 1.59 (1.16 – 2.18) | 1.98 (1.64 – 2.39) |
| Non-invasive ventilation use |  | P < 0.001 | P = 0.0069 | P = 0.0059 |
|  | No | 1.0 (ref) | 1.0 (ref) | 1.0 (ref) |
|  | Yes | 2.62 (2.34 – 2.93) | 4.02 (1.47 – 11) | 2.85 (1.35 – 6) |

Results from the sensitivity analyses, limited to patients ever admitted to ICU, can be found in Appendix 7, Appendix 8 and Appendix 9. The proportion of patients who ever were admitted to ICU was lower in the second wave than in the first wave (Table 2). During the second wave, compared to the first wave, ICU patients had a longer time from diagnosis to first ICU admission (median 4 days IQR 0 - 8 days, versus median 3 days IQR 0 – 6 days), shorter time from first ICU admission to outcome (median 18 days IQR 10 – 32 days, versus median 23 days IQR 12 - 41 days), a significantly lower use of invasive ventilation (60.7% versus 82.9%), and an increased risk of death (HR 1.39, 95% CI 1.32 – 1.45).

## Discussion

Based on CH-SUR data, we investigated the changes of COVID-19 in-hospital mortality over time in Switzerland, accounting for well-known risk factors for mortality such as age, sex or comorbidities. In-hospital mortality was lower and the time from diagnosis to death was longer during the second wave of COVID-19 compared to the first wave.

In contrast, in-IMCU/ICU mortality significantly increased in the second wave compared to the first wave. Time from diagnosis to first IMCU/ICU admission was longer in the second wave compared to the first wave. Time from first IMCU/ICU admission to outcome appeared to be similar between the first and the second wave, whereas time from first ICU admission to outcome appeared to be shorter in the second wave. The use of invasive ventilation was significantly less frequent in the second wave, whereas the use of non-invasive ventilation increased from the first to the second wave.

The lower in-hospital mortality observed in CH-SUR for the second wave seems not to be explained by changes in demographic characteristics of patients, since patients during the second wave tend to be slightly older, more comorbid, and they were more often male (all three being known to increase the risk of COVID-19 related death [6–9]). However, the changes in case management over time, induced by a growing knowledge on COVID-19, probably played a role in reducing in-hospital mortality. The use of corticosteroids (e.g. Dexamethasone), which is recommended systematically since summer 2020 by WHO for patients with severe or critical Covid-19 [18], and which was widely used during the second wave, has been shown to reduce mortality among patients receiving oxygen or invasive ventilation [19].

Moreover, a significantly lower referral rate of patients with a poor prognosis in the second wave could also have contributed to the reduction of observed in-hospital mortality over time while increasing the out-of-hospital mortality. Such a phenomenon could concern typically over-80-year-old persons who would have been hospitalised during the first wave but who were not because of their serious prognosis and the heaviness of acute care (either according to their will or due to triage [20]).

Compared to neighbouring countries in Western Europe, Switzerland had a lower in-hospital mortality (first wave 15.6%, second wave 14.0%) than e.g. France (first wave 16.2%, second wave 17.7%) [21] or Germany (first wave 19.1%, second wave 19.8%) [22], during both waves. This is a probable consequence of the good general health condition in Switzerland (reflected by one of the highest life expectancy worldwide [23]), a country with a well-developed health system, where comorbidities known as risk factors for COVID-19 mortality (e.g. cardiovascular diseases) are carefully controlled.

However, Switzerland was less successful at containing the pandemic during the second wave than other European countries. Its overall mortality (e.g number of deaths per million, see Appendix 10) and excess mortality [24] both increased during the second wave, despite being among the lowest in Europe during the first wave. This discrepancy between the time trends of in-hospital mortality and overall mortality related to COVID-19 further supports the hypothesis of increased out-of-hospital mortality during the second wave.

The characteristics of IMCU/ICU and ICU patients in CH-SUR during the first and second wave revealed significant changes in the case management and in the course of hospitalisation of COVID-19 patients. Those observations are in line with the updated guidelines in Swiss hospitals regarding acute care of hospitalised COVID-19 patients [25–28]. After the first COVID-19 wave, admission in ICU was avoided or postponed as much as possible, limiting it to the most severe cases, with a wider use of intermediate care facilities in hospitals having such a unit. Moreover, invasive ventilation, which was used for most ICU patients during the first wave, was partly replaced by non-invasive ventilation or high-flow nasal cannula [29] when possible, to avoid the risks associated with mechanical ventilation, including bacterial or fungal co-infections [30]).

The higher in-IMCU/ICU mortality observed in CH-SUR for the second wave could be a consequence of the high burden of COVID-19 cases on Swiss hospitals in the early weeks of the second wave, up to the saturation of ICU capacity in several cantons (particularly in the Western part of Switzerland) in November 2020. After that date, when the epidemic was on exponential increase, NPI where re-introduced, whereas the stringency index was relatively low in preceding weeks compared to neighbouring western European countries [31, 32]. Alternatively, given the longer time elapsed between diagnosis and first IMCU/ICU admission in the second wave, patients admitted to ICU might have been in a more severe condition during the second wave, subsequently leading to worse outcomes, possibly due to delayed optimal care.

To our knowledge, this is one of the first studies investigating the time trends of COVID-19 related in-hospital mortality in Switzerland. Using CH-SUR data allowed us to perform analyses including clinical information on comorbidities and ensuring a high coverage of COVID-19 hospitalisations in Switzerland, with about 67% of all hospitalisations reported mandatorily to FOPH by May 15^th^ 2021 included in CH-SUR. By imputing missing values of covariables, accounting for competing risks, and including age as a continuous variable (instead of categorizing it arbitrarily), we improved the accuracy of our approach. In contrast, as our study used observational patient data, it may be subject to collider bias [33].

Other studies in Europe have investigated the risk of death among patients infected by some SARS-CoV-2 Variants of Concern (VOCs) [34], e.g. the Alpha variant which appears to increase the risk of critical care admission and mortality compared to the common SARS-CoV-2 strain [35, 36], or the Delta variant which was shown to double the risk of hospital admission compared to Alpha variant [37]. The emergence of several suspected more virulent variants since Fall 2020 likely influences the time trends of COVID-19 related mortality. Information on the variant type has been recorded in the CH-SUR database since January 2021 for a fraction of documented episodes. However, to date data remains very sparse, which limits analyses on the effect of variants on COVID-19 in-hospital mortality.

Vaccination should also affect significantly trends in COVID-19 mortality, by reducing the risk of developing a severe form of the disease, and therefore reducing the risk of hospitalisation and death. In Switzerland, vaccination campaign started in early January 2021, first limited to very specific subgroups of the population. Consequently, we do not expect that it played a role in the differences in mortality between the first and the second wave. Nevertheless, vaccination will be a key element of analyses on COVID-19 related morbidity and mortality for the subsequent waves.

In conclusion, we found that in Switzerland COVID-19 in-hospital mortality was lower during the second wave compared to the first wave. In contrast, in the second wave, mortality significantly increased among patients admitted to intermediate or intensive care, compared to the first wave. We put our findings in perspective with the changes operated between the two time periods in terms of case management, treatment strategy, hospital burden and NPI. Further analyses in the near future, including more data from the last period, and taking into account the potential effect of variants and vaccination on mortality, would be crucial to have a complete overview of COVID-19 mortality trends throughout the different phases of the pandemic.

## Data Availability

The anonymised data can be accessed through a multi-stage process described elsewhere [11]. Applicants must fill a concept-sheet and send it to the team in charge of the study. An Executive Committee of experts and representatives of hospital participants will review the concept. Depending on the goal of the analysis, additional ethics clearance might be needed. Data will be restricted to the request and shared through a secure platform.

## Acknowledgements

The authors would like to thank all the participating centres’ teams, study nurses, and physicians for their hard work and commitment to the study. The Clinical Research Centre (Geneva University Hospitals and Faculty of Medicine) hosted the database.

## Ethical Approval

This study was submitted and approved by the Geneva Ethics Committee (CCER) and by all hospitals’ local Ethics Committees through the Swiss ethics BASEC submission system, under reference 2020-00827.

## Appendix

**Appendix 1:**
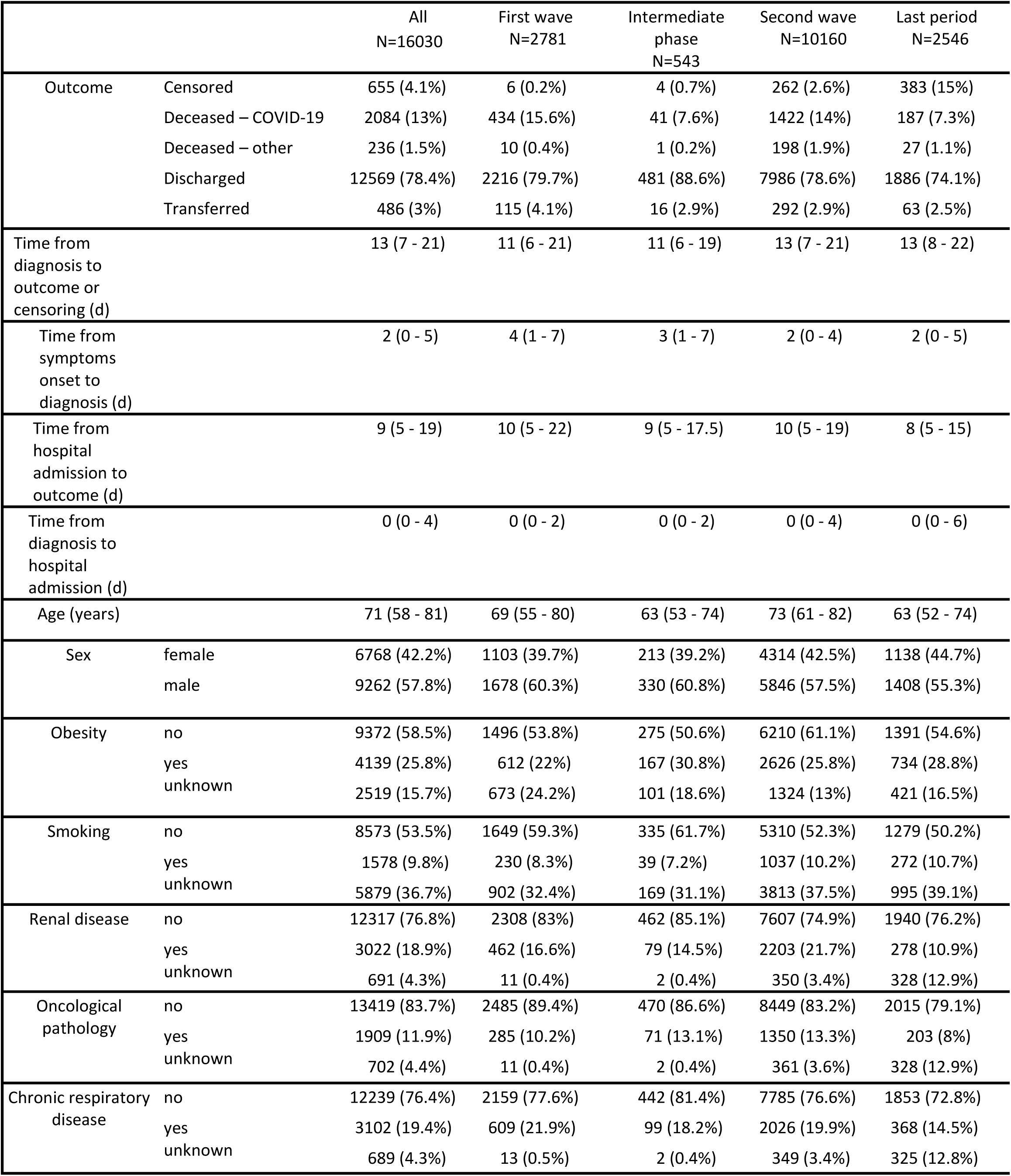
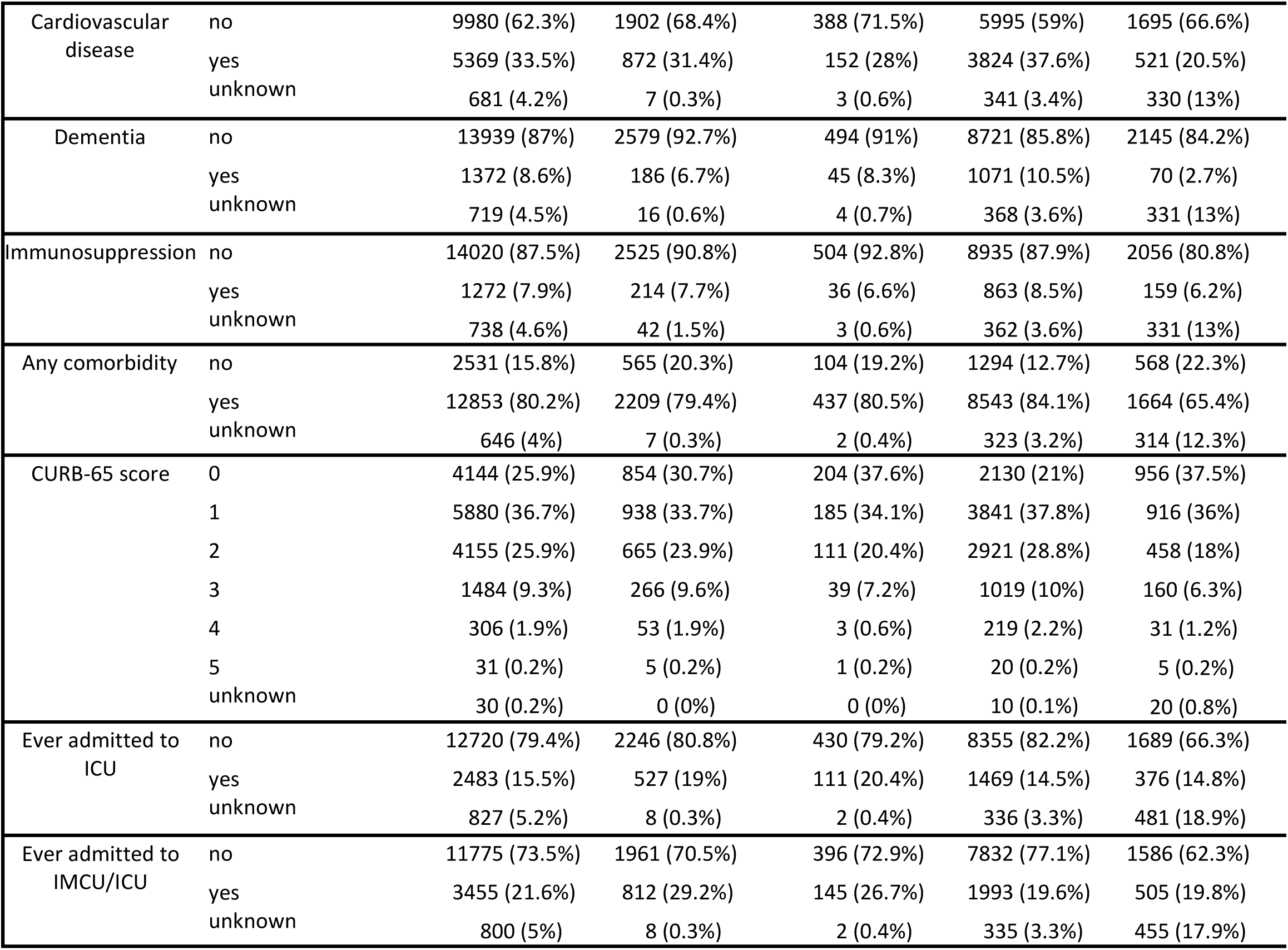
Characteristics of all COVID-19 episodes.

**Appendix 2:**
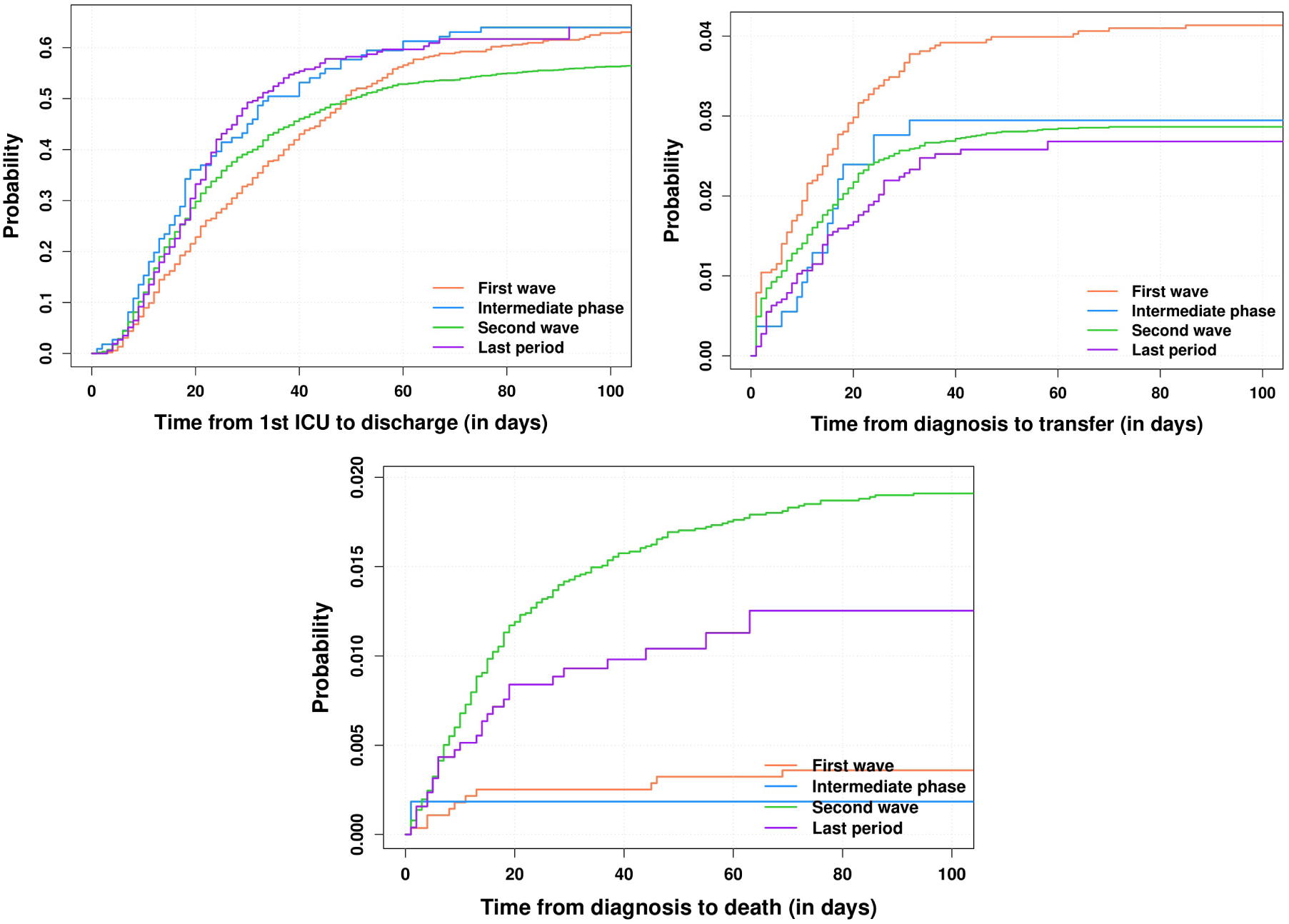
Cumulative incidence functions for discharge (upper left), transfer (upper right) and death due to other cause than COVID-19 (bottom) for the waves of COVID-19.

**Appendix 3:**
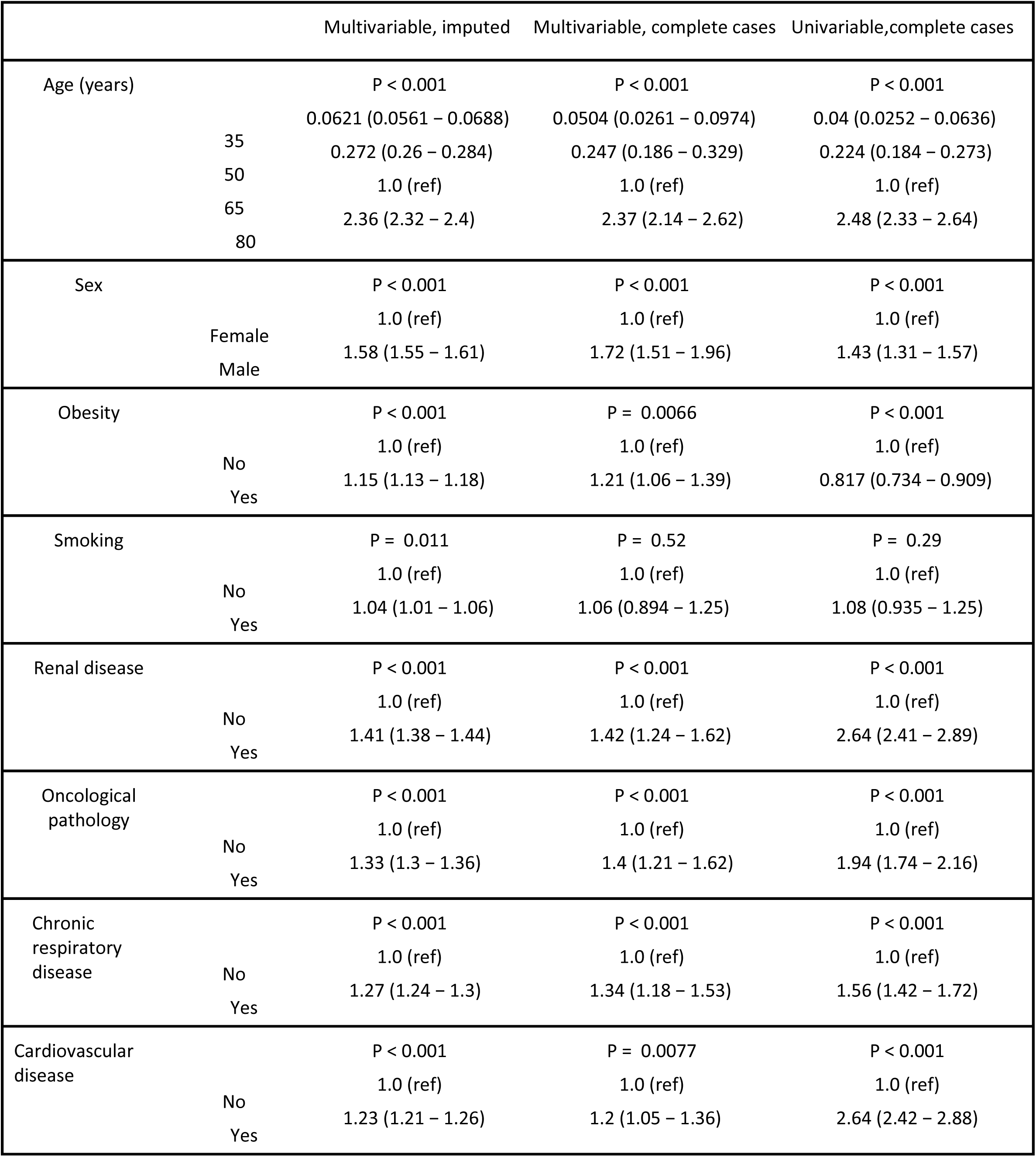
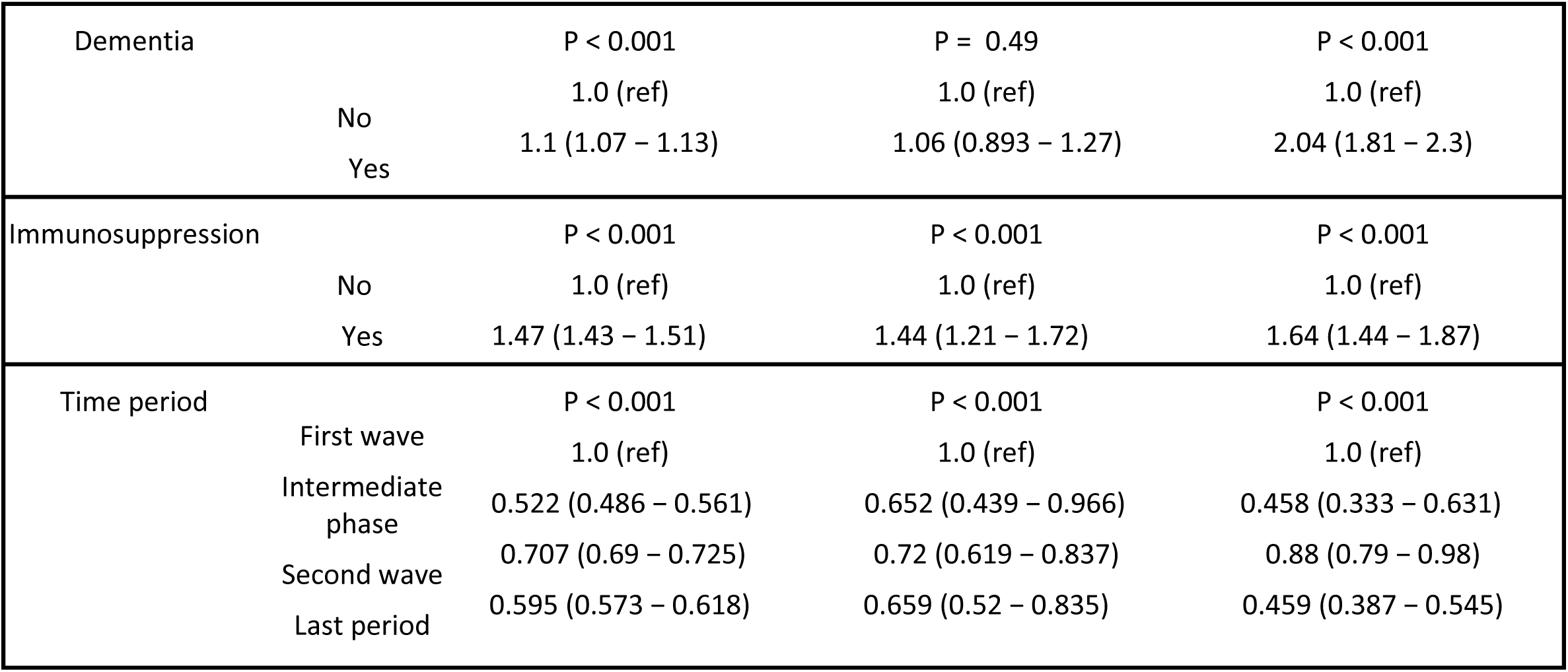
Hazard ratios from the multivariable Fine and Gray model with imputation of missing variables, multivariable Fine and Gray model on complete cases, and univariable Fine and Gray model on complete cases.

**Appendix 4:** Characteristics of episodes for patients who ever were admitted to IMCU/ICU.

|  |  | All<br>N=3429 | First wave<br>N=811 | Intermediate<br>phase<br>N=144 | Second wave<br>N=1980 | Last period<br>N=494 |
| --- | --- | --- | --- | --- | --- | --- |
| Outcome | Censored | 127 (3.7%) | 4 (0.5%) | 3 (2.1%) | 56 (2.8%) | 64 (13%) |
|  | Deceased - COVID-19 | 814 (23.7%) | 154 (19%) | 23 (16%) | 536 (27.1%) | 101 (20.4%) |
|  | Deceased – other | 75 (2.2%) | 7 (0.9%) | 1 (0.7%) | 57 (2.9%) | 10 (2%) |
|  | Discharged | 2169 (63.3%) | 580 (71.5%) | 101 (70.1%) | 1213 (61.3%) | 275 (55.7%) |
|  | Transferred | 244 (7.1%) | 66 (8.1%) | 16 (11.1%) | 118 (6%) | 44 (8.9%) |
| Time from first IMCU/ICU admission to outcome or censoring (d) |  | 16 (8 - 30) | 16 (8 - 34) | 18 (10 - 32) | 16 (9 - 30) | 17 (9 - 27) |
| Time from diagnosis to first IMCU/ICU admission (d) |  | 3 (0 - 7) | 2 (0 - 5) | 1 (0 - 3) | 4 (0 - 8) | 4 (0 - 8) |
| Age (years) |  | 68 (58 - 76) | 66 (56 - 75) | 64 (57.8 - 72.2) | 70 (60 - 77) | 65 (56 - 73) |
| Sex | female | 1103 (32.2%) | 244 (30.1%) | 48 (33.3%) | 624 (31.5%) | 187 (37.9%) |
|  | male | 2326 (67.8%) | 567 (69.9%) | 96 (66.7%) | 1356 (68.5%) | 307 (62.1%) |
| Obesity | no | 1762 (51.4%) | 375 (46.2%) | 62 (43.1%) | 1091 (55.1%) | 234 (47.4%) |
|  | yes | 1081 (31.5%) | 217 (26.8%) | 51 (35.4%) | 631 (31.9%) | 182 (36.8%) |
|  | unknown | 586 (17.1%) | 219 (27%) | 31 (21.5%) | 258 (13%) | 78 (15.8%) |
| Smoking | no | 1833 (53.5%) | 469 (57.8%) | 77 (53.5%) | 1025 (51.8%) | 262 (53%) |
|  | yes | 414 (12.1%) | 82 (10.1%) | 12 (8.3%) | 252 (12.7%) | 68 (13.8%) |
|  | unknown | 1182 (34.5%) | 260 (32.1%) | 55 (38.2%) | 703 (35.5%) | 164 (33.2%) |
| Renal disease | no | 2827 (82.4%) | 688 (84.8%) | 122 (84.7%) | 1591 (80.4%) | 426 (86.2%) |
|  | yes | 589 (17.2%) | 120 (14.8%) | 22 (15.3%) | 383 (19.3%) | 64 (13%) |
|  | unknown | 13 (0.4%) | 3 (0.4%) | 0 (0%) | 6 (0.3%) | 4 (0.8%) |
| Oncological pathology | no | 3003 (87.6%) | 733 (90.4%) | 127 (88.2%) | 1688 (85.3%) | 455 (92.1%) |
|  | yes | 412 (12%) | 76 (9.4%) | 17 (11.8%) | 283 (14.3%) | 36 (7.3%) |
|  | unknown | 14 (0.4%) | 2 (0.2%) | 0 (0%) | 9 (0.5%) | 3 (0.6%) |
| Chronic respiratory disease | no | 2601 (75.9%) | 612 (75.5%) | 116 (80.6%) | 1466 (74%) | 407 (82.4%) |
|  | yes | 813 (23.7%) | 197 (24.3%) | 28 (19.4%) | 507 (25.6%) | 81 (16.4%) |
|  | unknown | 15 (0.4%) | 2 (0.2%) | 0 (0%) | 7 (0.4%) | 6 (1.2%) |
| Cardiovascular disease | no | 2215 (64.6%) | 566 (69.8%) | 100 (69.4%) | 1207 (61%) | 342 (69.2%) |
|  | yes | 1205 (35.1%) | 245 (30.2%) | 44 (30.6%) | 769 (38.8%) | 147 (29.8%) |
|  | unknown | 9 (0.3%) | 0 (0%) | 0 (0%) | 4 (0.2%) | 5 (1%) |
| Dementia | no | 3292 (96%) | 786 (96.9%) | 138 (95.8%) | 1888 (95.4%) | 480 (97.2%) |
|  | yes | 127 (3.7%) | 24 (3%) | 6 (4.2%) | 87 (4.4%) | 10 (2%) |
|  | unknown | 10 (0.3%) | 1 (0.1%) | 0 (0%) | 5 (0.3%) | 4 (0.8%) |
| Invasive ventilation | no | 837 (24.4%) | 91 (11.2%) | 36 (25%) | 559 (28.2%) | 151 (30.6%) |
|  | yes | 1612 (47%) | 435 (53.6%) | 75 (52.1%) | 886 (44.7%) | 216 (43.7%) |
|  | unknown | 980 (28.6%) | 285 (35.1%) | 33 (22.9%) | 535 (27%) | 127 (25.7%) |
| Non-invasive ventilation | no | 71 (2.1%) | 17 (2.1%) | 1 (0.7%) | 48 (2.4%) | 5 (1%) |
|  | yes | 2020 (58.9%) | 436 (53.8%) | 75 (52.1%) | 1188 (60%) | 321 (65%) |
|  | unknown | 1338 (39%) | 358 (44.1%) | 68 (47.2%) | 744 (37.6%) | 168 (34%) |
| Immunosuppression | no | 3087 (90%) | 738 (91%) | 135 (93.8%) | 1767 (89.2%) | 447 (90.5%) |
|  | yes | 312 (9.1%) | 62 (7.6%) | 9 (6.2%) | 201 (10.2%) | 40 (8.1%) |
|  | unknown | 30 (0.9%) | 11 (1.4%) | 0 (0%) | 12 (0.6%) | 7 (1.4%) |
| Any comorbidity | no | 500 (14.6%) | 144 (17.8%) | 23 (16%) | 229 (11.6%) | 104 (21.1%) |
|  | yes | 2927 (85.4%) | 667 (82.2%) | 121 (84%) | 1751 (88.4%) | 388 (78.5%) |
|  | unknown | 2 (0.1%) | 0 (0%) | 0 (0%) | 0 (0%) | 2 (0.4%) |
| IMCU/ICU type | both | 510 (14.9%) | 179 (22.1%) | 17 (11.8%) | 268 (13.5%) | 46 (9.3%) |
|  | ICU only | 1952 (56.9%) | 345 (42.5%) | 94 (65.3%) | 1190 (60.1%) | 323 (65.4%) |
|  | IMCU only | 937 (27.3%) | 285 (35.1%) | 33 (22.9%) | 520 (26.3%) | 99 (20%) |
|  | unknown | 30 (0.9%) | 2 (0.2%) | 0 (0%) | 2 (0.1%) | 26 (5.3%) |
| CURB-65 score | 0 | 693 (20.2%) | 204 (25.2%) | 34 (23.6%) | 328 (16.6%) | 127 (25.7%) |
|  | 1 | 1203 (35.1%) | 287 (35.4%) | 52 (36.1%) | 658 (33.2%) | 206 (41.7%) |
|  | 2 | 1012 (29.5%) | 197 (24.3%) | 40 (27.8%) | 657 (33.2%) | 118 (23.9%) |
|  | 3 | 409 (11.9%) | 96 (11.8%) | 17 (11.8%) | 262 (13.2%) | 34 (6.9%) |
|  | 4 | 94 (2.7%) | 25 (3.1%) | 0 (0%) | 61 (3.1%) | 8 (1.6%) |
|  | 5 | 18 (0.5%) | 2 (0.2%) | 1 (0.7%) | 14 (0.7%) | 1 (0.2%) |
|  | unknown | 0 (0%) | 0 (0%) | 0 (0%) | 0 (0%) | 0 (0%) |

**Appendix 5:**
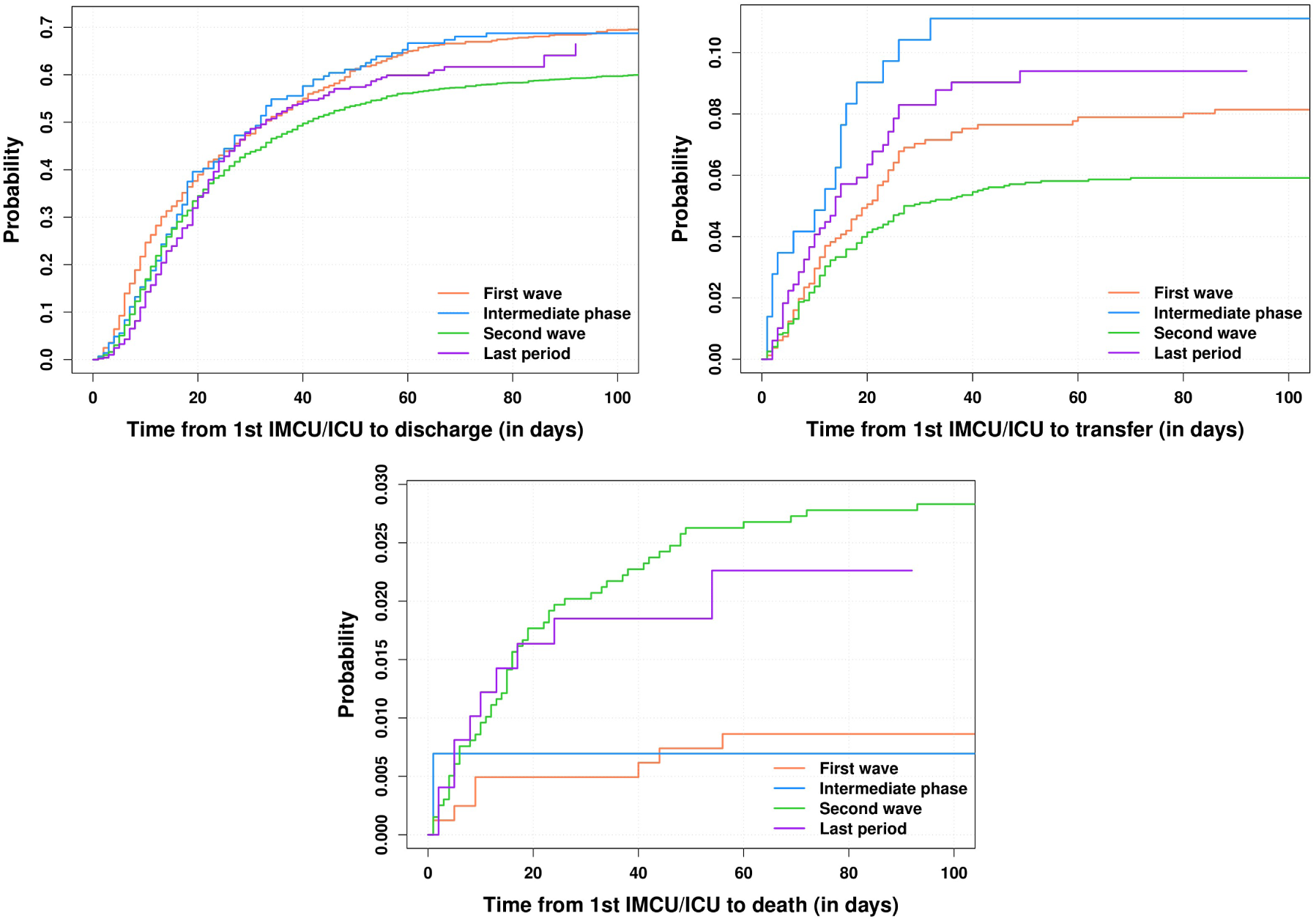
Cumulative incidence functions for discharge (upper left), transfer (upper right) and death from other cause than COVID-19 (bottom), for IMCU/ICU patients and for the different COVID-19 waves.

**Appendix 6:**
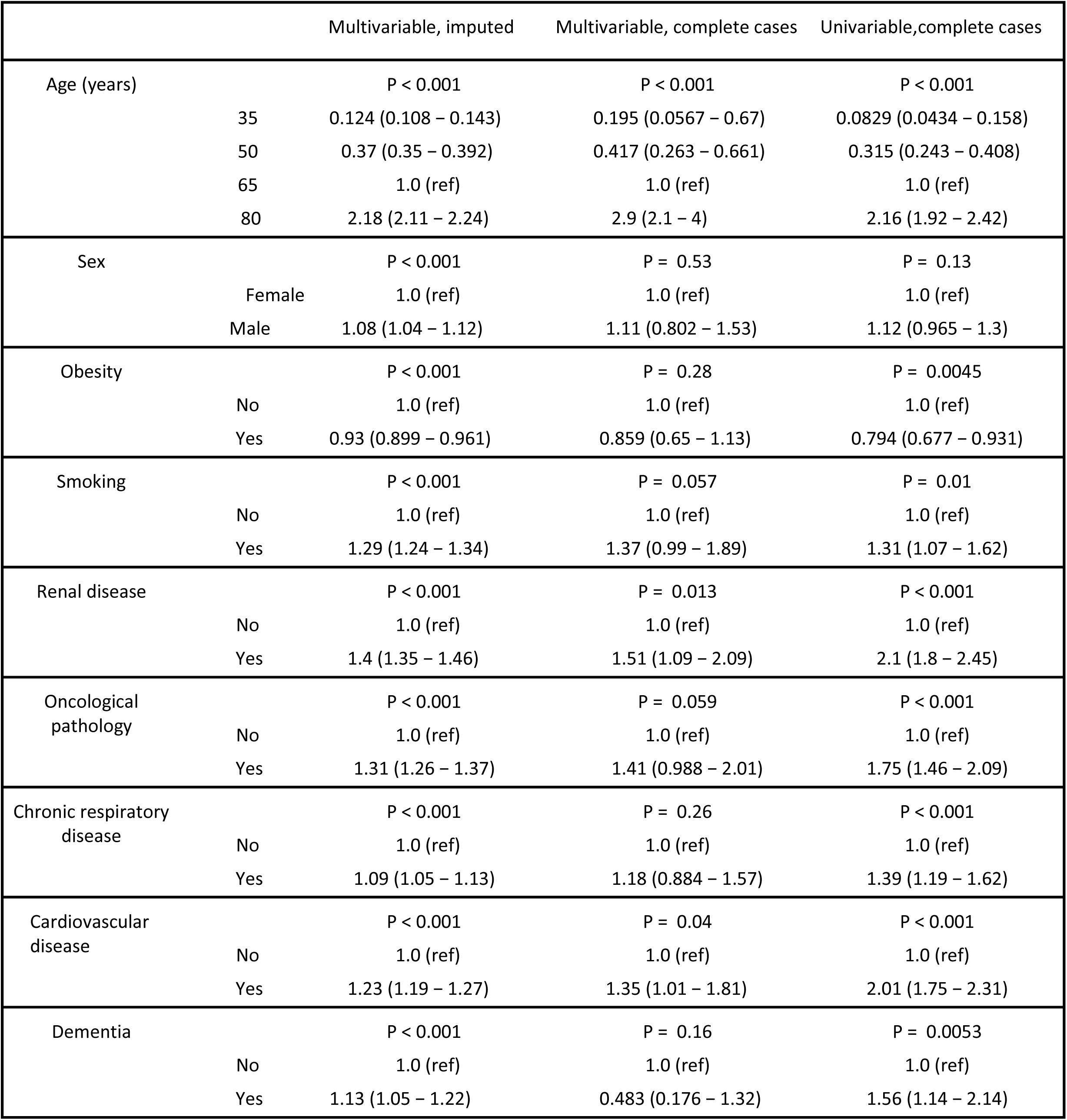
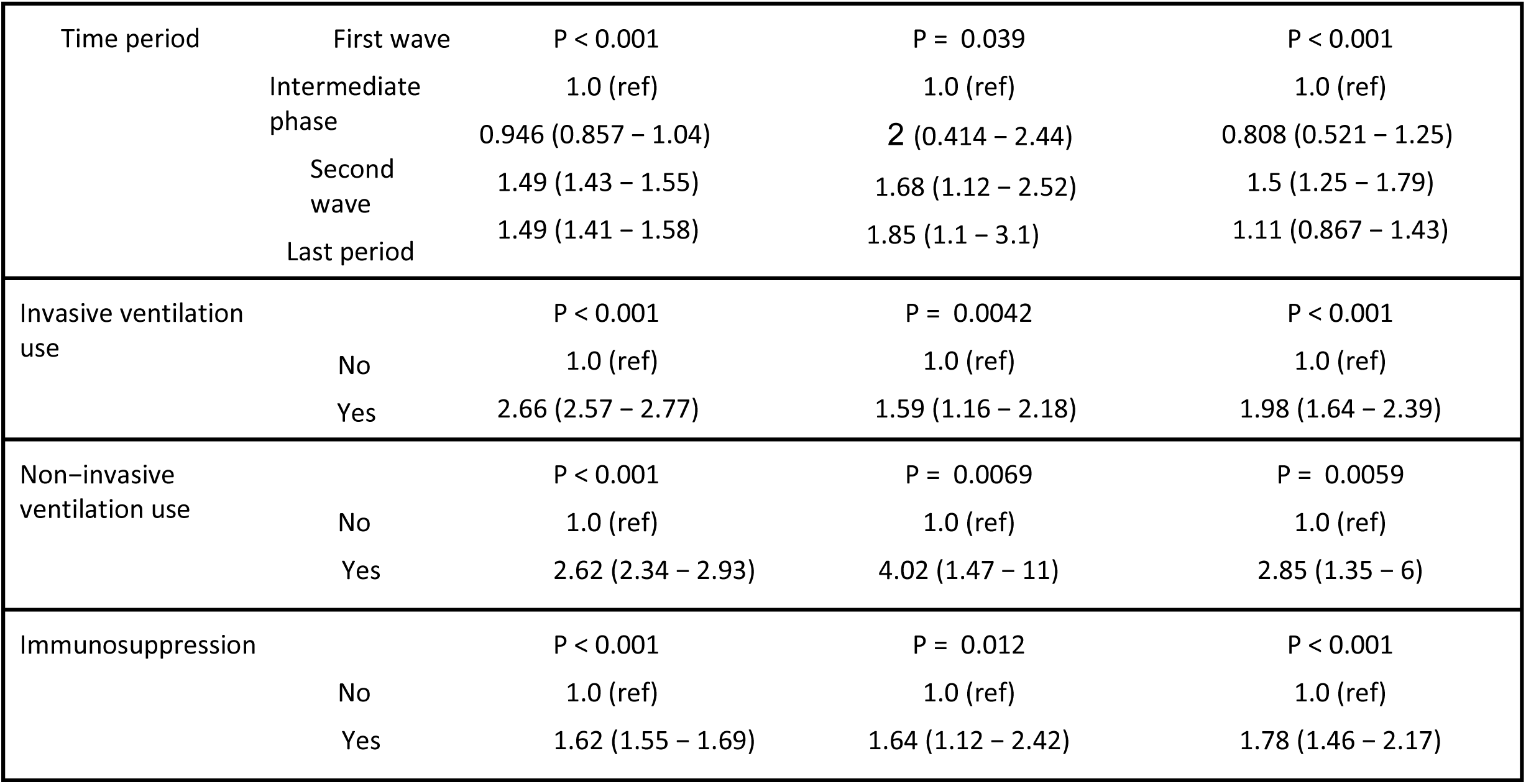
Hazard ratios for in-hospital mortality from the various Fine-Gray models calculated, only patients ever admitted to IMCU/ICU.

**Appendix 7:** Characteristics of episodes for patients who ever were admitted to ICU.

|  |  | All<br>N=2466 | First wave<br>N=525 | Intermediate phase<br>N=111 | Second wave<br>N=1458 | Last period<br>N=372 |
| --- | --- | --- | --- | --- | --- | --- |
| Outcome | Censored | 91 (3.7%) | 2 (0.4%) | 3 (2.7%) | 51 (3.5%) | 35 (9.4%) |
|  | Deceased - COVID-19 | 641 (26%) | 120 (22.9%) | 18 (16.2%) | 426 (29.2%) | 77 (20.7%) |
|  | Deceased - other | 54 (2.2%) | 6 (1.1%) | 1 (0.9%) | 39 (2.7%) | 8 (2.2%) |
|  | Discharged | 1467 (59.5%) | 344 (65.5%) | 73 (65.8%) | 840 (57.6%) | 210 (56.5%) |
|  | Transferred | 213 (8.6%) | 53 (10.1%) | 16 (14.4%) | 102 (7%) | 42 (11.3%) |
| Time from first ICU admission to outcome or censoring (d) |  | 19 (10 - 33) | 23 (12 - 41) | 18 (10 - 32) | 18 (10 - 32) | 18 (10 - 26) |
| Time from diagnosis to first ICU admission (d) |  | 3 (0 - 7) | 3 (0 - 6) | 0 (0 - 3) | 4 (0 - 8) | 4 (0 - 8) |
| Age (years) |  | 67 (58 - 74) | 66 (57 - 74) | 63 (57 - 70) | 68 (59 - 75) | 64 (56 - 71) |
| Sex | female | 734 (29.8%) | 131 (25%) | 36 (32.4%) | 436 (29.9%) | 131 (35.2%) |
|  | male | 1732 (70.2%) | 394 (75%) | 75 (67.6%) | 1022 (70.1%) | 241 (64.8%) |
| Obesity | no | 1243 (50.4%) | 261 (49.7%) | 44 (39.6%) | 770 (52.8%) | 168 (45.2%) |
|  | yes | 844 (34.2%) | 155 (29.5%) | 44 (39.6%) | 495 (34%) | 150 (40.3%) |
|  | unknown | 379 (15.4%) | 109 (20.8%) | 23 (20.7%) | 193 (13.2%) | 54 (14.5%) |
| Smoking | no | 1232 (50%) | 290 (55.2%) | 59 (53.2%) | 705 (48.4%) | 178 (47.8%) |
|  | yes | 305 (12.4%) | 48 (9.1%) | 9 (8.1%) | 192 (13.2%) | 56 (15.1%) |
|  | unknown | 929 (37.7%) | 187 (35.6%) | 43 (38.7%) | 561 (38.5%) | 138 (37.1%) |
| Renal disease | no | 2042 (82.8%) | 454 (86.5%) | 93 (83.8%) | 1174 (80.5%) | 321 (86.3%) |
|  | yes | 412 (16.7%) | 69 (13.1%) | 18 (16.2%) | 278 (19.1%) | 47 (12.6%) |
|  | unknown | 12 (0.5%) | 2 (0.4%) | 0 (0%) | 6 (0.4%) | 4 (1.1%) |
| Oncological pathology | no | 2187 (88.7%) | 485 (92.4%) | 96 (86.5%) | 1261 (86.5%) | 345 (92.7%) |
|  | yes | 268 (10.9%) | 40 (7.6%) | 15 (13.5%) | 189 (13%) | 24 (6.5%) |
|  | unknown | 11 (0.4%) | 0 (0%) | 0 (0%) | 8 (0.5%) | 3 (0.8%) |
| Chronic respiratory disease | no | 1889 (76.6%) | 400 (76.2%) | 94 (84.7%) | 1087 (74.6%) | 308 (82.8%) |
|  | yes | 565 (22.9%) | 124 (23.6%) | 17 (15.3%) | 365 (25%) | 59 (15.9%) |
|  | unknown | 12 (0.5%) | 1 (0.2%) | 0 (0%) | 6 (0.4%) | 5 (1.3%) |
| Cardiovascular disease | no | 1633 (66.2%) | 363 (69.1%) | 80 (72.1%) | 919 (63%) | 271 (72.8%) |
|  | yes | 824 (33.4%) | 162 (30.9%) | 31 (27.9%) | 535 (36.7%) | 96 (25.8%) |
|  | unknown | 9 (0.4%) | 0 (0%) | 0 (0%) | 4 (0.3%) | 5 (1.3%) |
| Dementia | no | 2388 (96.8%) | 514 (97.9%) | 111 (100%) | 1399 (96%) | 364 (97.8%) |
|  | yes | 71 (2.9%) | 11 (2.1%) | 0 (0%) | 56 (3.8%) | 4 (1.1%) |
|  | unknown | 7 (0.3%) | 0 (0%) | 0 (0%) | 3 (0.2%) | 4 (1.1%) |
| Invasive ventilation | no | 836 (33.9%) | 90 (17.1%) | 36 (32.4%) | 559 (38.3%) | 151 (40.6%) |
|  | yes | 1611 (65.3%) | 435 (82.9%) | 75 (67.6%) | 885 (60.7%) | 216 (58.1%) |
|  | unknown | 19 (0.8%) | 0 (0%) | 0 (0%) | 14 (1%) | 5 (1.3%) |
| Immunosuppression | no | 2211 (89.7%) | 474 (90.3%) | 106 (95.5%) | 1297 (89%) | 334 (89.8%) |
|  | yes | 232 (9.4%) | 41 (7.8%) | 5 (4.5%) | 154 (10.6%) | 32 (8.6%) |
|  | unknown | 23 (0.9%) | 10 (1.9%) | 0 (0%) | 7 (0.5%) | 6 (1.6%) |
| Any comorbidity | no | 369 (15%) | 94 (17.9%) | 17 (15.3%) | 174 (11.9%) | 84 (22.6%) |
|  | yes | 2095 (85%) | 431 (82.1%) | 94 (84.7%) | 1284 (88.1%) | 286 (76.9%) |
|  | unknown | 2 (0.1%) | 0 (0%) | 0 (0%) | 0 (0%) | 2 (0.5%) |
| CURB-65 score | 0 | 475 (19.3%) | 107 (20.4%) | 26 (23.4%) | 243 (16.7%) | 99 (26.6%) |
|  | 1 | 871 (35.3%) | 195 (37.1%) | 40 (36%) | 476 (32.6%) | 160 (43%) |
|  | 2 | 721 (29.2%) | 139 (26.5%) | 31 (27.9%) | 470 (32.2%) | 81 (21.8%) |
|  | 3 | 305 (12.4%) | 62 (11.8%) | 13 (11.7%) | 205 (14.1%) | 25 (6.7%) |
|  | 4 | 78 (3.2%) | 20 (3.8%) | 0 (0%) | 52 (3.6%) | 6 (1.6%) |
|  | 5 | 16 (0.6%) | 2 (0.4%) | 1 (0.9%) | 12 (0.8%) | 1 (0.3%) |
|  | unknown | 0 (0%) | 0 (0%) | 0 (0%) | 0 (0%) | 0 (0%) |

**Appendix 8:**
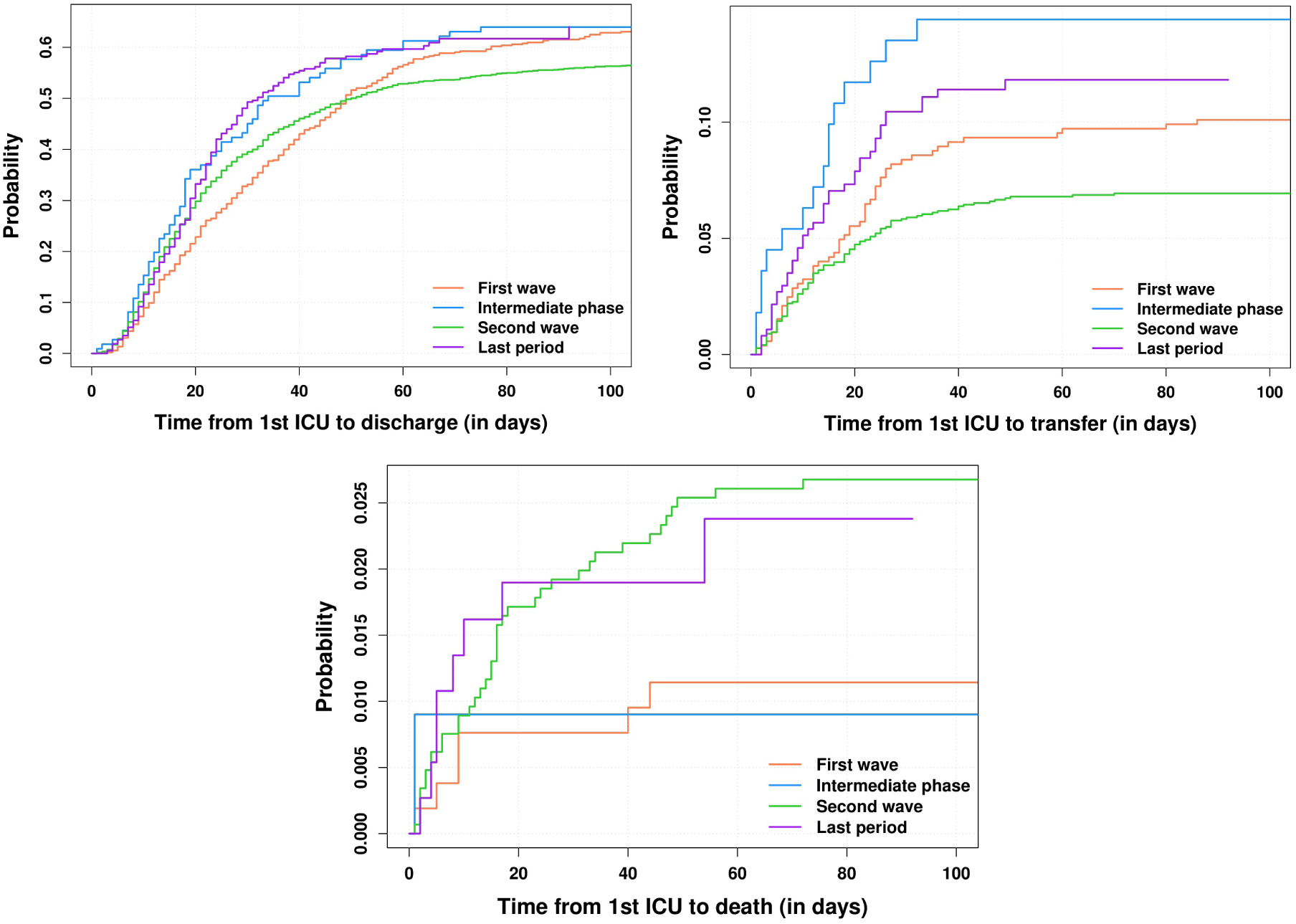
Cumulative incidence functions for discharge (upper left), transfer (upper right) and death from other cause than COVID-19 (bottom), for ICU patients and for the different COVID-19 waves.

**Appendix 9:**
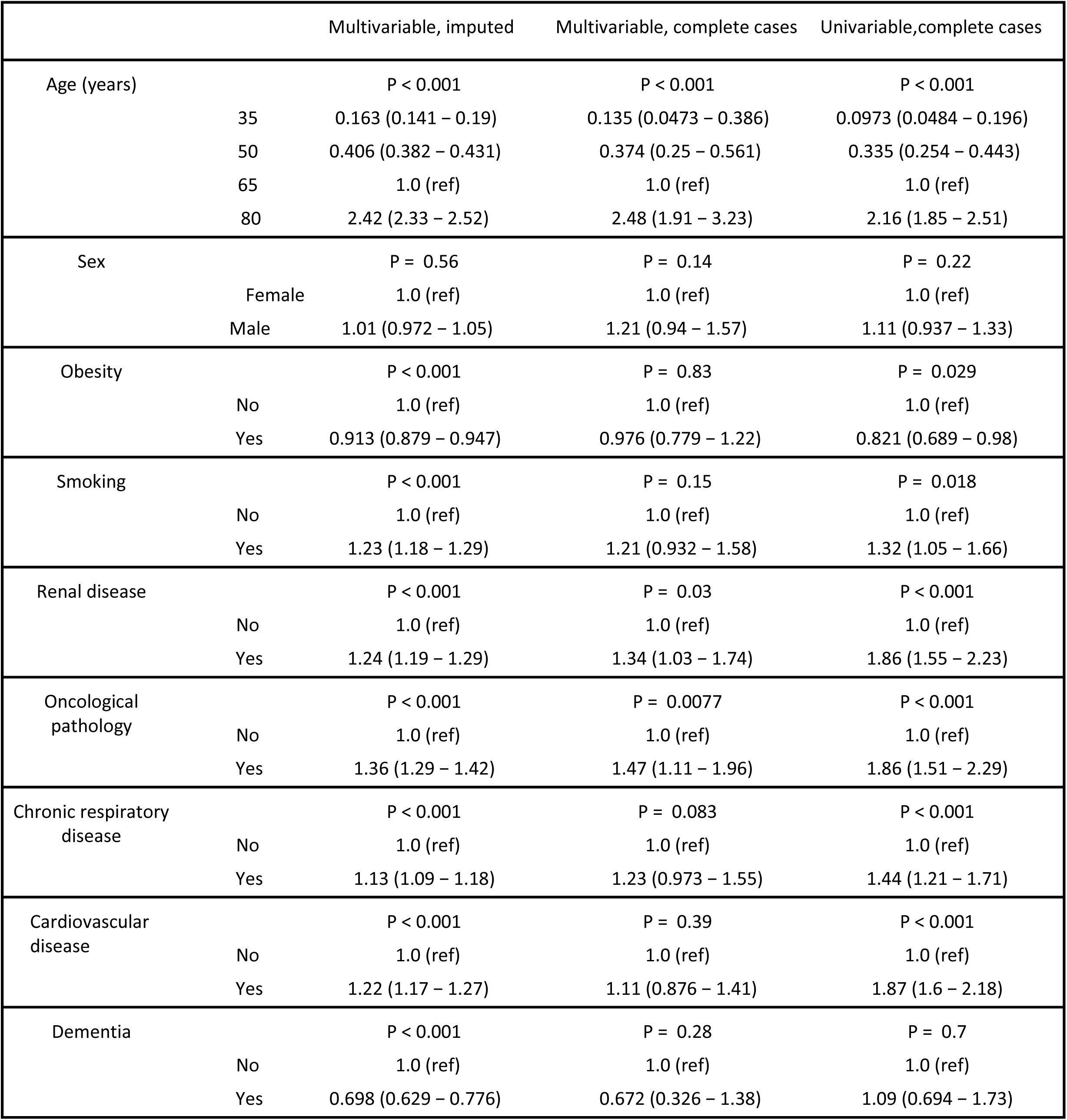
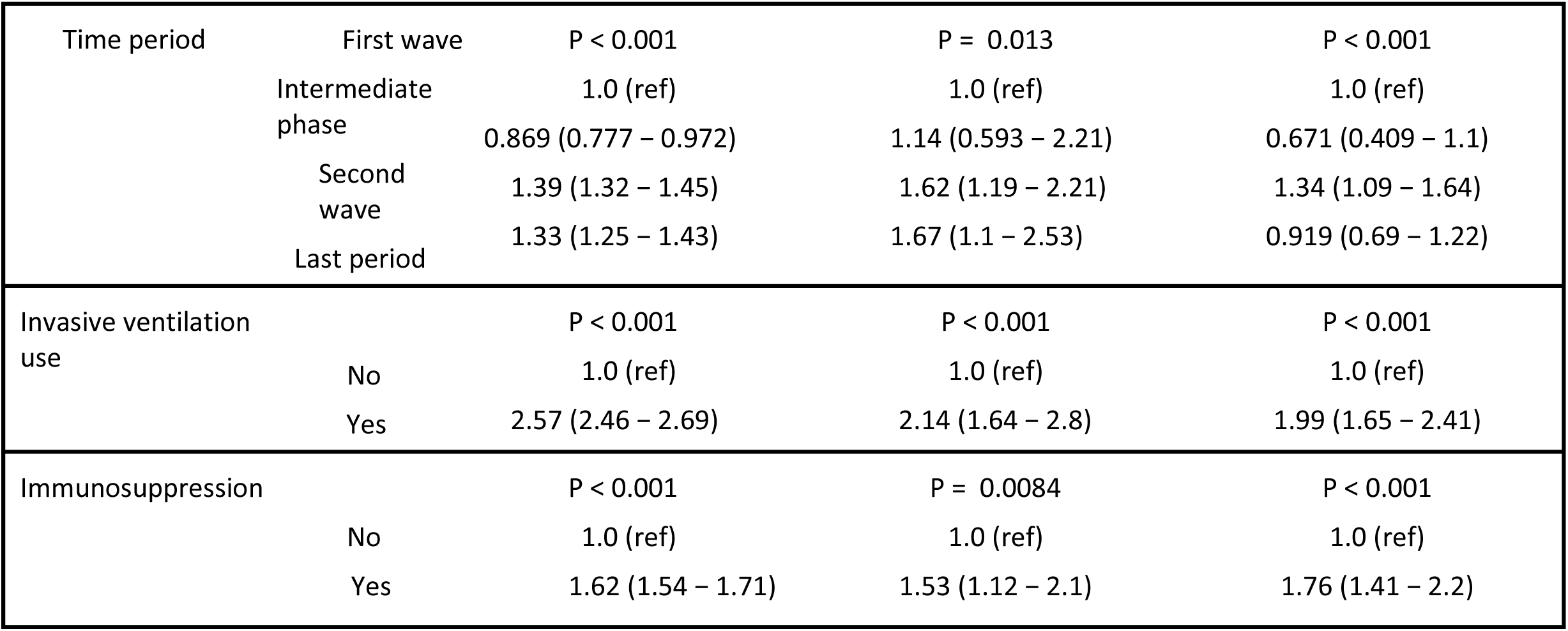
Hazard ratios for in-hospital mortality from the various Fine-Gray models calculated, only patients ever admitted to ICU.

**Appendix 10:**
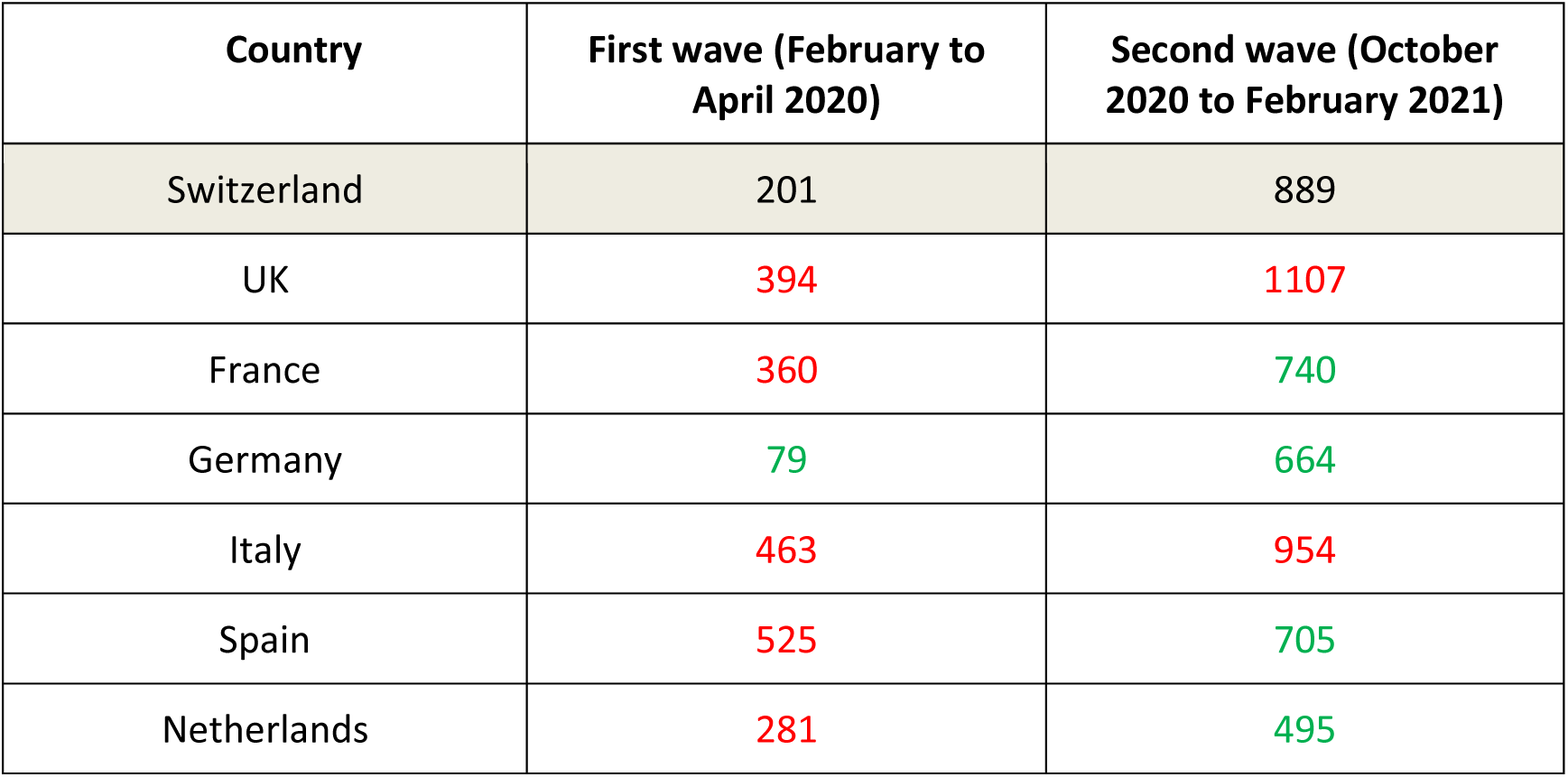
COVID-19 mortality in some European countries, expressed in deaths per million, from Our World in Data (OWID) [25]. Red colour indicates numbers higher than in Switzerland for the period of interest, green colour indicates numbers lower than Switzerland for the period of interest.

